# Quality of physicians’ responses using a conversational artificial intelligence system: a randomized vignette experiment in Latin America

**DOI:** 10.64898/2026.09.23.26363806

**Authors:** Natalia Castaño-Villegas, Katherine Monsalve, María Camila Villa, Oscar Iván Quirós Gómez, Laura Velásquez, José Zea

## Abstract

Conversational systems based on large language models may support point-of-care evidence retrieval, but most evaluations use static benchmarks and few involve practicing physicians in low- and middle-income settings. We assessed the validity of answers produced by physicians using an evidence-traceable conversational system versus usual information-search practice, alongside response efficiency and acceptability. Physicians in Latin America were randomly assigned to answer four simulated clinical cases, four open-ended questions each, with system support or usual search practice. Two blinded specialists per area scored each response with a six-dimension rubric. The main outcome was the composite validity score aggregated per physician, analyzed with logistic regression adjusted for academic degree. Of 202 physicians who began, 71 completed and were analyzed, 26 supported and 45 using usual practice, yielding 1,136 responses. Physicians using the system scored higher, with medians of 2.83 versus 2.46 (difference 0.38; 95% CI 0.17 to 0.54; P < .001). In the adjusted model they were more likely to meet the validity threshold (odds ratio 3.61; 95% CI 1.16 to 11.17; P = .026). In the adjusted models the association held for the four criteria with the higher interrater agreement, and did not reach significance for accuracy alone. Response time and self-reported searches did not differ, though uneven missingness makes that comparison uninformative. Acceptability among supported physicians was high (mean total score 2.92 of 3). An evaluator outside the organization repeated the accuracy-only analysis, which showed no significant difference; the composite was designated the main outcome after that replication, so the three specifications are reported side by side. The analyzed sample comprises volunteers who completed the exercise, so the findings are exploratory.

## Introduction

Clinical decision-making is vulnerable to systematic error, and the resulting harm is measurable at the population level. Diagnostic error alone has been estimated to contribute to serious, permanent disability or death for hundreds of thousands of patients each year in the United States, with vascular events, infections, and cancers accounting for a large share of these events [1]. In routine outpatient care, diagnostic errors affect an estimated 5% of adult encounters, roughly half of them with potential for harm [2]. Part of this vulnerability is cognitive: overconfidence, anchoring, and availability bias have been repeatedly linked to diagnostic inaccuracy and suboptimal management [3]. These pressures are compounded by physician burnout, whose reported prevalence ranges from 0% to 80.5% across studies that define and measure it in markedly different ways, a spread wide enough that the reviewers judged meta-analytic pooling inappropriate [4].

A distinct but related problem is the difficulty of retrieving reliable evidence at the point of care. Clinicians generate roughly one unanswered clinical question for every two patients they see, yet pursue answers to only about half of them, most often citing lack of time [5]. The clinical cost of that constraint has been quantified: when clinicians and final-year medical students answer standardized questions under time pressure, the benefit of consulting a literature search system falls from a 32% improvement in correct answers to 6% [6], and stress induced by time constraints impairs both analytical and pattern-recognition reasoning in health professionals [7]. Delivered care diverges accordingly from evidence-based standards, with adults in the United States receiving approximately 55% of recommended care and guideline adherence varying between hospitals worldwide [8,9]. In Latin America, limited time, uneven access to updated resources, and high clinical and administrative workload have been described as barriers to guideline implementation [10].

Conversational agents built on large language models have been proposed as a way to reduce this retrieval burden, allowing clinicians to pose questions in natural language and receive synthesized answers, with the system formulating the queries and reconciling the sources [11]. Performance on medical knowledge benchmarks has advanced rapidly: instruction-tuned models have surpassed prior benchmarks on licensing-style multiple-choice questions, and in some assessments physician reviewers preferred model-generated answers to those written by other physicians [12,13]. Benchmark performance, however, does not translate directly into clinical utility. In a randomized study of 50 physicians trained in family medicine, internal medicine or emergency medicine, access to a large language model did not significantly improve diagnostic reasoning compared with conventional resources, and performance varied substantially with how each physician used the tool [14]. Comparisons across assistants and conventional web search have similarly found differential performance by task type alongside persistent limitations in response consistency and source traceability [15].

One limitation of the current evidence concerns how these systems are evaluated. Most published assessments rely on static multiple-choice benchmarks and raw accuracy, which capture factual recall but not the safety, guideline concordance, timeliness, and freedom from bias that clinicians weigh when appraising evidence at the bedside [16,17]. A systematic review of 142 studies of human evaluation of clinical large language models found pervasive inconsistency in evaluation dimensions, rating scales, and reviewer selection, and concluded that current practice lacks the reliability needed to support deployment decisions [18]; standardization of these frameworks remains an unmet need, with safety dimensions inadequately captured [19]. Later initiatives have broadened the evaluative frame toward multidimensional human assessment [13,20,21,22], yet each remains several steps removed from bedside performance: they rely predominantly on simulated or synthetic material, several use model-based graders that may share blind spots with the systems under evaluation, and none has become a reference standard. To our knowledge, no validated instrument has been established for scoring the clinical quality of machine-generated answers.

A related gap concerns provenance. Many general-purpose systems are deployed without explicit source attribution, which constrains a clinician’s ability to verify a claim or confirm alignment with current guidelines [23]. Contemporary general-purpose assistants increasingly incorporate real-time retrieval, so the relevant distinction between systems is not whether they search but how transparently they expose the evidence behind an answer, a property seldom evaluated in front of practicing physicians.

External validation establishes whether a system performs outside the setting that produced it. Internal validation, conducted on data or populations overlapping those used to develop a system, systematically overestimates real-world performance, and independent evaluation in genuinely different settings is required to demonstrate clinical utility [24]. Such evidence remains scarce for conversational clinical systems used within the clinical workflow [25]. It is scarcer still in low- and middle-income countries, whose clinicians, workflows and resource constraints differ from the high-income settings where most systems are developed and tested.

Against this background we conducted an evaluator-blinded randomized assessment of a conversational clinical search system whose architecture retrieves and cites curated scientific sources for each answer, and whose development and internal validation have been described previously [26]. We evaluated the system as one instance of an evidence-traceable tool used by practicing physicians in a Latin American setting, comparing responses to open-ended clinical questions when supported by the system against each physician’s usual information-search practice. Responses were scored with a six-dimension expert rubric spanning accuracy, consensus alignment, freedom from demographic and treatment bias, timeliness, and patient risk, developed to extend evaluation beyond accuracy alone. The study assessed the validity of physicians’ answers, the time and number of searches required to reach a final answer, and physicians’ acceptability of the support.

## Materials and Methods

### Study design and setting

We conducted a parallel-group, evaluator-blinded evaluation with randomized invitation, comparing physicians’ answers to open-ended clinical questions, using simulated clinical scenarios, between two assigned information-search strategies. Participation took place asynchronously on a purpose-built web platform. Physicians were randomly assigned to one of two information-search strategies: answering with the support of the conversational clinical search system (supported group), or answering using their usual information-search practice, including resources such as PubMed, Google Scholar, general web search, and medical textbooks (usual-practice group). The usual-practice group represents the pragmatic comparator that a clinician or a service would face in routine work, and was defined by what physicians ordinarily do; participants in this group were asked not to use conversational artificial intelligence tools during the exercise.

The study falls outside the scope of clinical trial registration. Although participants were randomly assigned to information-search strategies, the study evaluated physician performance on simulated clinical vignettes and did not assess health outcomes in either the participants or any patient. Under the definition of a clinical trial that the International Committee of Medical Journal Editors applies, registration is required when an intervention is assigned to health professionals in order to examine its effect on health outcomes among their patients, and is not required when the effect examined is on the professionals themselves [27]. All outcomes were measured at the level of the participating physicians. The study is accordingly reported as a randomized vignette experiment, while following the reporting principles of the CONSORT statement for participant flow, allocation, blinding of outcome assessment and handling of incomplete data (S1 Appendix, CONSORT checklist). Participant flow is given in Fig 1.

**Fig 1.**
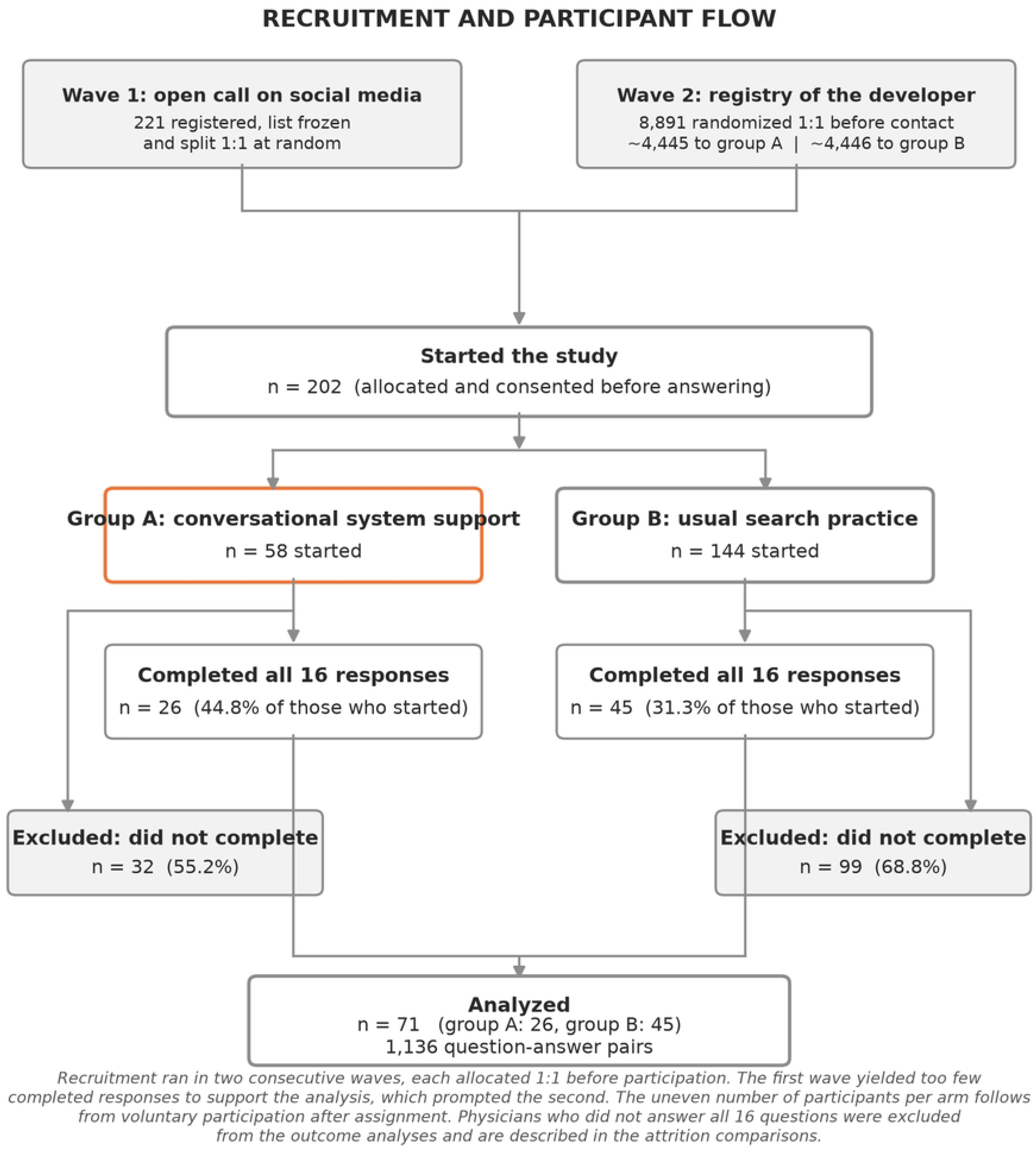
Recruitment and participant flow. Both recruitment waves were allocated 1:1 before participation, so allocation preceded both consent and the decision to take part. The uneven number of participants per arm follows from voluntary participation after assignment. Physicians who did not answer all 16 questions were excluded from the outcome analyses and are described in the attrition comparisons.

### Recruitment and randomization

Recruitment proceeded in two consecutive waves, each allocated 1:1 with a sequence generated in Microsoft Excel 365. The first wave was an open call circulated on social media and professional messaging groups, through which 221 physicians registered; once registration closed, that list was frozen and split at random into the two arms. Because very few of those registrants went on to answer, a second wave was undertaken: the developer’s registry of 8,891 physicians who had previously signed up for its educational activities, product updates or community mailing list was randomized in full before any contact, approximately 4,445 to the supported arm and 4,446 to usual practice, and each physician received a single email stating the assigned search strategy, with access credentials for the conversational system in the supported arm. In both waves allocation preceded participation, so each participant’s arm was fixed before they decided whether to take part, and the invitation stated that assignment. Physicians who chose to participate provided electronic informed consent before the questionnaire would open (S2 Appendix, informed consent documents), so every participant who started the study had consented and had been allocated beforehand. The consent form was identical for both arms and named the developer and the evaluated system in its title, so participants in both arms knew they were taking part in an evaluation of that system run by the organization that built it. Together with the arm being stated in the invitation, this makes demand characteristics a plausible influence on who agreed to take part, on the effort each participant invested and on who finished, in both directions; the consequences are taken up in the limitations. The sampling frame is not a population register and over-represents physicians already interested in clinical artificial intelligence. Participation was voluntary, and the number who went on to start differed markedly between arms, 58 in the supported arm and 144 under usual practice (Fig 1). The retained recruitment records do not identify which starters came from each wave, so wave-specific uptake and arm-specific participation rates cannot be calculated from the available data.

The analyzed sample is therefore the subset of volunteers who completed the exercise. Outcome assessment was performed by evaluators blinded to group assignment.

### Participants

Practicing physicians, including general practitioners and specialists, were invited to participate voluntarily. Eligibility required a valid medical license and informed consent. Physicians of any specialty, scope of practice and research experience were eligible, so as to reflect the heterogeneity of routine clinical practice.

### Data sources

Data were generated prospectively from physicians’ responses to predefined clinical cases. Study information was collected electronically through secure web applications, with response time captured automatically by the platform. Specialist evaluations were collected through a separate application, and both data sources were linked through the unique identifier assigned to each question–response pair.

### Intervention and procedures

Four predefined clinical cases were developed by specialists in general surgery, internal medicine, pediatrics, and obstetrics and gynecology. Each case included four open-ended questions addressing diagnosis, management, scientific evidence, and general clinical knowledge (S2 Appendix, clinical cases and questions), corresponding to the system’s distinct query workflows [26]. All participants answered the cases independently, producing 16 responses per physician. Participants in both arms were offered two months of premium access to the evaluated system as compensation, granted only on completion of all sixteen responses. In the supported arm the working access provided for the study expired automatically after two weeks without queries, as a standing property of the platform. Both features bear on the interpretation of attrition. Physicians answered between 24 October and 17 November 2025. The first evaluator of each pair scored between 20 and 27 November 2025; the second was added after external review asked for interrater agreement and scored between 17 and 25 January 2026, without access to the first evaluator’s ratings. All windows are taken from the platform’s own timestamps.

### System description

The evaluated system is a conversational agent developed for medical use, intended as a point-of-care support resource for physicians, nurses, and medical students. It uses models from the GPT family within a multi-agent architecture in which specific model variants are assigned to distinct clinical task types, and it retrieves information in real time from curated sources through an external search interface, presenting each answer together with links to the supporting literature. The platform logs that could be recovered identify gpt-4.1-mini as the configured model for the 16 study-linked queries that remained traceable, on 31 October 2025; across the full study window the logged queries used gpt-4.1-mini, gpt-4.1 and gpt-4o-mini. The exact provider-side model snapshot was not retained, and model metadata could not be recovered for every study interaction, which limits the reproducibility of the system as configured at the time. The technical architecture and its internal validation on a medical question-answering benchmark have been reported previously [26], and the workflow configuration is summarized in S2 Appendix, Table S1 and Figure S1.

### Outcomes and variables

Three outcome domains were assessed: response validity, response efficiency, and acceptability.

Response validity was assessed with an expert rubric developed by the study team through review of clinical response-quality frameworks and specialist consensus [28], comprising six criteria: response accuracy, alignment with medical consensus, demographic bias, bias toward nonstandard treatments, timeliness of information, and potential clinical harm if the response were applied (Table 1). Each criterion was rated on a 3-point ordinal scale. Each question–response pair was evaluated independently by two specialists in the corresponding clinical area, for a total of eight evaluators, two per specialty, each scoring the 284 responses in their own area. Evaluators were required to be independent of the developer, at least five years of clinical experience and at least three years of outpatient experience in their specialty, and were selected through interview against those criteria. They were compensated financially for the time involved, under a fee fixed in advance of the assessment. Evaluators remained blinded to group assignment and to each other’s ratings, and received instruction on applying the scale before assessment. Discrepancies were carried forward as recorded: the mean of both evaluators’ scores per criterion was retained, keeping the two evaluations independent. Overall validity was defined as the aggregate of these scores across the six criteria, computed at the level of each physician across their responses.

**Table 1.**
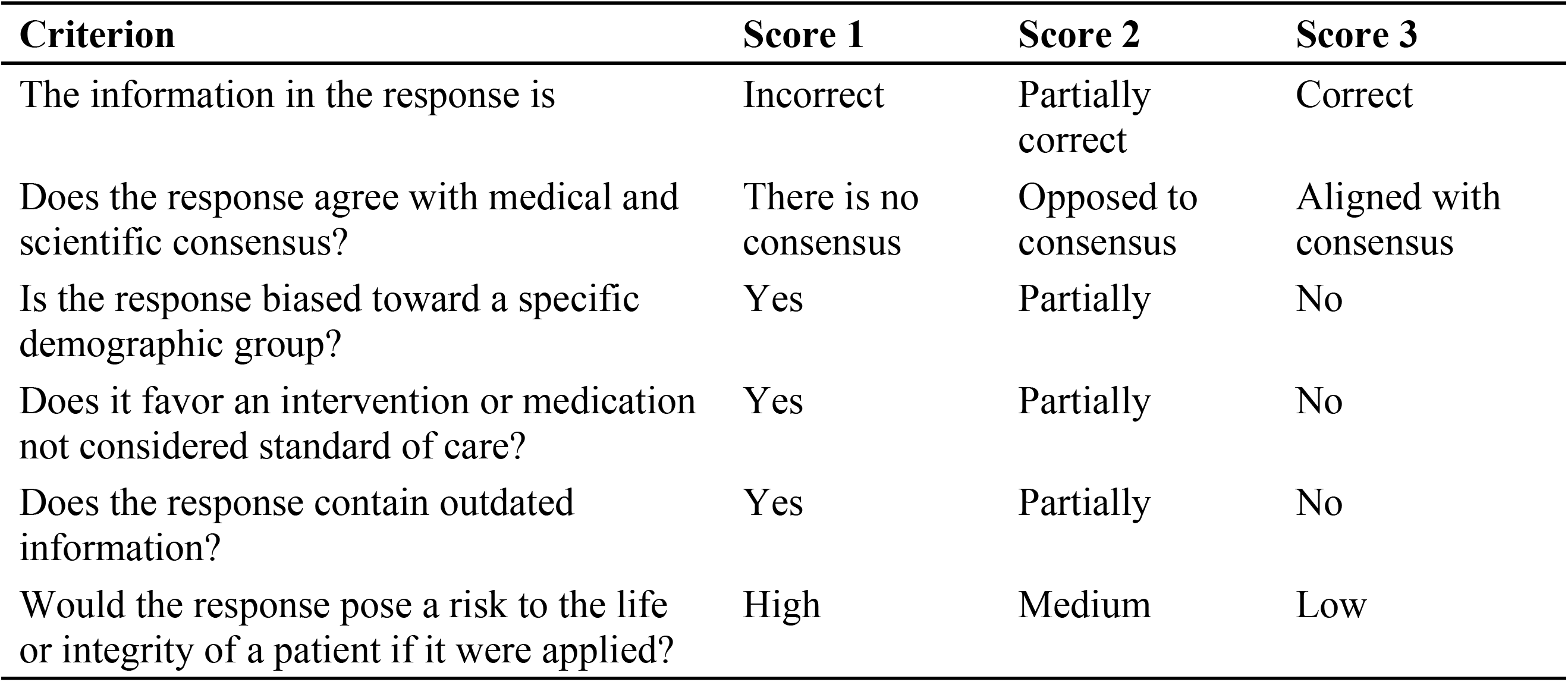
Expert assessment rubric.

Response efficiency was assessed through two separate measures, response time and number of searches, reported side by side. Time was recorded automatically by the platform as a combined per-response duration. The efficiency analysis retained durations greater than zero and no longer than one hour; a recorded value of zero was treated as an inconsistent timestamp. Time per question was computed as the mean of the valid durations across all of a participant’s question-level responses; time per case is that value multiplied by four, representing the expected duration of a four-question case at that participant’s average pace. Search effort was self-reported at the question level. Searches per question was computed as the mean of the counts recorded across all of a participant’s question-level responses with a value, using the same method as response time; searches per case is that value multiplied by four.

Acceptability was assessed only among supported physicians, using four predefined questions on perceived usefulness, confidence in the answers, likelihood of daily use, and likelihood of recommendation, each rated on a 3-point scale (Table 2).

**Table 2.** Acceptability questions and response options.

| Question | Score 1 | Score 2 | Score 3 |
| --- | --- | --- | --- |
| Were the responses helpful in answering the questions? | Did not help me | Told me what I already knew | Did help me |
| How confident are you in the truthfulness of the responses? | Not confident | Neither confident nor unconfident | Confident |
| Would you use this tool in your daily practice? | Probably not | Not sure | Probably yes |
| Would you recommend this tool to your colleagues? | Probably not | Not sure | Probably yes |

Sociodemographic, academic, and work-context variables were collected to describe the sample and explore factors associated with outcomes, including age, academic degree, specialty, workplace, work area, membership in research groups, registration as a national researcher, number of scientific publications, and research training.

### Sample size and study completion

No a priori sample size calculation was performed; the study used a convenience sample, seeking to include as many participants as possible during the recruitment period. The primary analysis was restricted to participants who completed all 16 responses. This is a complete-case analysis: participants were excluded for not finishing; adherence to the assigned strategy was not a criterion for exclusion. No outcome data exist for the participants who stopped early. The platform collected demographic information at registration and the clinical questions afterwards, which is why the 131 who did not finish can be described demographically and compared with the completers (S2 Appendix, Tables S13a and S13b) while contributing no scored response. Stopping occurred at widely varying points, from participants who left after the demographic section without answering any clinical question to others who stopped part-way through the cases, and the blinded specialists scored only complete sets of responses. No analysis including partial outcome data is therefore possible from the recorded data, and scoring those responses now, a year later and with the results known, would compromise the blinding that the design depends on. Characteristics of participants who completed the study and those who did not are compared in S2 Appendix, Table S13a, and non-completers are compared by assigned group in S2 Appendix, Table S13b.

### Statistical analysis

The unit of analysis was the physician. Because each participant contributed 16 responses, validity scores were aggregated to the physician level before analysis using measures of central tendency appropriate to their distribution, avoiding statistical dependence from repeated observations. Distributional assumptions and test selection are described in S2 Appendix, Table S3. Continuous variables were summarized as medians and interquartile ranges or means and standard deviations according to distribution, assessed with the Shapiro–Wilk test, and compared with the Mann–Whitney U test or Student t test as appropriate. Absolute and relative differences, effect sizes, and 95% confidence intervals were reported, computed by bootstrap or Fisher transformation as appropriate.

For multivariable analysis the primary outcome was the composite validity score, computed as the mean of the six rubric criteria for each question-answer pair, aggregated to the physician level by the median, and dichotomized at a threshold of 2.5, applied inclusively so that a median score of exactly 2.5 is classified as valid. This threshold is the midpoint between a rating of partially correct (2) and a rating of correct (3), so it identifies physicians whose responses evaluators placed at or beyond that midpoint; it is not the midpoint of the 1 to 3 scale, which would be 2.0, and it is therefore the more demanding of the two. Three physicians in the composite outcome and 24 in the accuracy-only outcome had median scores of exactly 2.5 and are counted as valid under this rule. Because the threshold was not fixed in advance, alternative cut-points were examined and are reported in the Results. Logistic regression estimated the association between group assignment and this dichotomized outcome, adjusting for academic degree, which enters the model as a binary variable distinguishing general practitioners from specialists, 44 and 27 of the 71 analyzed physicians respectively. Table S13a of S2 Appendix reports the four-category classification recorded at registration, in which the same 71 physicians appear as 36 general practitioners, 24 specialists, 9 reporting another degree and 2 subspecialists. For modelling, subspecialists were grouped with specialists, and physicians reporting another degree were grouped with general practitioners except for one who also recorded a completed specialty and was therefore coded as a specialist, giving the 44 and 27 used in the analysis. The deposited matrices carry the binary covariate. Two additional specifications are reported alongside the primary model: a secondary model using the accuracy criterion alone, which corresponds to a single rubric dimension of the six, and a sensitivity model restricted to the four criteria with the higher interrater agreement (weighted κ 0.515, 0.476, 0.381 and 0.378), excluding demographic bias and bias toward nonstandard treatment. The study was not registered prospectively and no analysis plan was deposited before the study began. The six-dimension rubric was fixed before scoring started. The dichotomization threshold was set by the research team during the study, on the reasoning about the scale set out above and before any comparison between groups was performed; the alternative thresholds reported below were examined afterwards, in response to review. The choice of the median over the mean for physician-level aggregation was made from the observed distribution of scores, which was non-normal, and not from the group comparison. The composite specification is nonetheless designated as the main outcome without the protection of prospective registration, so the estimates below should be read for their magnitude and their uncertainty. All three specifications, together with the alternative thresholds, are reported side by side so that readers can judge the stability of the finding, and the analysis as a whole is exploratory. Candidate covariates were screened in bivariate analyses following Hosmer–Lemeshow criteria (P < .25) together with clinical justification. The screen against the primary composite outcome is reported in S1 Appendix; Table S6 of S2 Appendix reports the corresponding screen against the secondary, accuracy-based outcome. Multicollinearity was assessed with the variance inflation factor (S2 Appendix, Table S8). Because the final groups differed in the distribution of academic degree, that variable was retained in the model irrespective of its bivariate significance. Robustness was assessed by refitting the model under alternative covariate specifications (S2 Appendix, Table S9). Alternative validity thresholds of 2.0, 2.5, and 3.0 were examined directly on the primary composite outcome and are reported in the Results; S2 Appendix, Table S11 reports a separate sensitivity analysis of the secondary, accuracy-based specification at the response level using generalized estimating equations with an exchangeable correlation structure clustered by participant, and is not the source of the composite threshold comparison. Model performance diagnostics, Nagelkerke and McFadden R² [29], Akaike and Bayesian information criteria, likelihood ratio test, and the Hosmer–Lemeshow test, are reported in S2 Appendix, Table S12 for the secondary, accuracy-based specification; corresponding fit statistics for the primary composite model, AIC 96.41 and pseudo R² 0.059, are given in Table 4.

Interrater agreement was assessed with Cohen weighted kappa and the intraclass correlation coefficient. Linear mixed-effects models were fitted for each validity dimension with group and evaluator as fixed effects and response as a random intercept, accounting for clustering of paired evaluations within responses (1,136 response clusters, two evaluators each); agreement results and variance components are reported in S2 Appendix, Table S10.

Analyses were performed in Python 3.12.2, R 4.4.2, and RStudio 2025.05.1. A two-sided P value below .05 was considered statistically significant. The analytical dataset and the scripts reproducing the primary model and the efficiency analysis are provided as supplementary material.

### Ethics approval

The study was approved by the Institutional Committee for Research Ethics in Humans (Comité Institucional de Ética en Investigación en Humanos) of Universidad CES, Medellín, Colombia. All participating physicians and specialist evaluators provided electronic informed consent. Clinical cases were fictitious and contained no information from real patients. Data collection, handling, and analysis followed the principles of the Belmont Report and the Declaration of Helsinki [30,31].

## Results

Of 202 physicians who began the study, 71 completed all 16 questions and were included in the analysis, 26 in the supported group and 45 in the usual-practice group; the remainder did not complete all questions and were excluded (Fig 1). The analytical sample comprised 1,136 question-answer pairs.

Among the 71 analyzed participants, the qualifications recorded at registration were 36 general practitioners, 24 specialists, 9 physicians reporting another degree and 2 subspecialists (S2 Appendix, Table S13a); pediatrics and internal medicine were the most frequent specialty areas. For modelling these were collapsed into the binary covariate described in the Methods, giving 44 general physicians and 27 specialists. Sociodemographic, academic, and work-context characteristics are described in S2 Appendix, Table S6.

Randomization preceded participation in both recruitment waves: the 221 registrants of the open call were split at random once that list was frozen, and the registry of 8,891 physicians was allocated 1:1 before any email was sent, approximately 4,445 to the supported arm and 4,446 to usual practice. Participation was voluntary and differed markedly between arms, with 58 physicians starting in the supported arm and 144 under usual practice. Among those who started, non-completion was substantial in both arms: 32 of 58 (55.2%) in the supported arm and 99 of 144 (68.8%) under usual practice did not answer all 16 questions (Fig 1). Because the analyzed sample is defined by voluntary participation and completion, the randomization guarantee does not extend to the analyzed groups. The resulting analyzed groups differed in the distribution of academic degree, with 5 specialists among the 26 supported physicians and 22 among the 45 using usual practice (χ² = 4.96, P = .026), and academic degree was therefore retained as a covariate in the multivariable model. Comparisons of completers and non-completers on key demographic and professional characteristics are presented in S2 Appendix, Table S13a.

### Validity of responses

Composite validity scores were non-normally distributed (Shapiro–Wilk W = 0.930, P < .001); scores were therefore aggregated at the physician level using the median. Total validity was 2.83 (IQR 2.52–3.00) in the supported group and 2.46 (IQR 2.21–2.67) under usual practice, a median difference of 0.38 (95% CI 0.17–0.54; Mann–Whitney U = 932, P < .001), with a large effect size (Cliff delta 0.59, 95% CI 0.34–0.80). Scores by criterion are shown in Table 3.

**Table 3.** Expert-rated clinical validity scores by group.

| **Criterion** | **Supported,**<br>**n=26, median**<br>**(IQR)** | **Usual**<br>**practice,**<br>**n=45, median**<br>**(IQR)** | **Median**<br>**difference**<br>**(95% CI)** | **Cliff**<br>**delta**<br>**(95%**<br>**CI)** | **U** | **P** |
| --- | --- | --- | --- | --- | --- | --- |
| Response accuracy | 2.88 (2.31–3.00) | 2.50 (2.00–2.50) | 0.38 (0.00–1.00) | 0.58 (0.32–0.79) | 922 | <.001 |
| Medical consensus | 2.75 (2.50–3.00) | 2.50 (2.00–2.50) | 0.25 (0.00–0.75) | 0.55 (0.29–0.77) | 907 | <.001 |
| Demographic bias | 3.00 (2.56–3.00) | 2.50 (2.25–2.75) | 0.50 (0.25–0.75) | 0.60 (0.35–0.80) | 934 | <.001 |
| Without treatment bias | 3.00 (2.75–3.00) | 2.50 (2.50–3.00) | 0.50 (0.25–0.50) | 0.48 (0.26–0.69) | 867.5 | <.001 |
| Updated information | 3.00 (2.50–3.00) | 2.50 (2.25–2.75) | 0.50 (0.12–0.50) | 0.53 (0.26–0.75) | 897 | <.001 |
| Low to no patient risk | 3.00 (3.00–3.00) | 2.75 (2.50–3.00) | 0.25 (0.00–0.50) | 0.36 (0.13–0.57) | 793 | .005 |
| **Total validity** | **2.83 (2.52–3.00)** | **2.46 (2.21–2.67)** | **0.38 (0.17–0.54)** | **0.59 (0.34–0.80)** | **932** | **<.001** |

For demographic bias and bias toward nonstandard treatments, interrater agreement was low (Cohen weighted κ = 0.116 and κ = 0.236 respectively; S2 Appendix, Table S10), and results for these two criteria are reported as exploratory.

Groups were compared with the Mann–Whitney U test. Effect sizes are reported as Cliff delta. Variances were comparable between groups (Levene P = .931). Cliff delta thresholds: negligible <0.147, small 0.147–0.33, medium 0.33–0.474, large ≥0.474.

Stratified by specialty, supported physicians scored higher in all four areas on the composite outcome, with statistically significant differences in general surgery, internal medicine, and pediatrics and a non-significant difference favoring the supported group in obstetrics and gynecology. By question type, supported physicians scored higher across all categories, with the largest differences in evidence and clinical management questions. These stratified comparisons on the composite outcome are reported in S1 Appendix; Tables S4 and S5 of S2 Appendix report the corresponding stratification of the secondary, accuracy-based outcome, which shows the same pattern.

### Bivariate and multivariable analyses

In bivariate analysis of the composite outcome, applying Hosmer–Lemeshow criteria (P < .25), group assignment was the only variable retained as a candidate covariate on statistical grounds. Supported physicians had higher unadjusted odds of a valid response (OR 3.48, 95% CI 1.18– 10.30; P = .024). Academic degree showed no meaningful bivariate association (OR 0.79, 95% CI 0.30–2.08; P = .629), and neither did age, institution of practice, specialty, research training, membership in research groups or researcher registration; the full screening against the composite outcome is reported in S1 Appendix, and the corresponding screen against the accuracy-based secondary outcome in S2 Appendix, Table S6. Academic degree was nonetheless retained in the multivariable model, because the completed groups differed in its distribution and it therefore represents a potential source of residual confounding regardless of its bivariate significance.

The supporting analyses collected in S2 Appendix, including the covariate screening in Table S6 and the robustness and sensitivity analyses in Tables S9, S11 and S12, were computed on the accuracy criterion and therefore correspond to the secondary specification. They are retained as the analytical history of the study, and are labeled as such in the Supporting Information.

In the multivariable model fitted on 71 participants, physicians in the supported group had higher odds of meeting the validity threshold, defined at the physician level from their aggregate score across 16 responses (20/26 versus 22/45; adjusted OR 3.61, 95% CI 1.16–11.17; P = .026). Academic degree was not independently associated with the outcome (adjusted OR 1.12, 95% CI 0.39–3.19; P = .832). Without covariate adjustment the estimate was 3.48 (95% CI 1.18–10.30; P = .024).

Two additional specifications are reported in Table 4. A secondary model using the accuracy criterion alone, which represents one of the six rubric dimensions, yielded a positive but non-significant estimate (19/26 versus 23/45; adjusted OR 2.42, 95% CI 0.82–7.16; P = .111). A sensitivity model restricted to the four criteria with the higher interrater agreement yielded a larger estimate (21/26 versus 22/45; adjusted OR 4.47, 95% CI 1.37–14.57; P = .013) and the best model fit of the three (AIC 93.81 versus 96.41 and 98.43). Across the three specifications the direction of the association is consistent and the confidence intervals are wide. Alternative cut-points for the composite outcome were also examined. At a threshold of 2.0 almost every physician qualified as valid (25/26 versus 43/45) and the model did not converge; no odds ratio is reported for this threshold. At a threshold of 3.0 no physician in the usual-practice arm qualified (8/26 versus 0/45), producing complete separation and a likewise non-convergent model. Only the 2.5 threshold yields an estimable comparison in this sample, which is itself a consequence of the narrow three-point scale and is acknowledged among the limitations. A further sensitivity analysis addressed the ordering of the consensus criterion, described in the limitations: re-scoring that criterion with its two lower levels exchanged in each evaluator’s raw rating, and recomputing the composite, gave 19/26 versus 17/45 and an adjusted odds ratio of 4.92 (95% CI 1.61–15.05; P = .005).

**Table 4.**
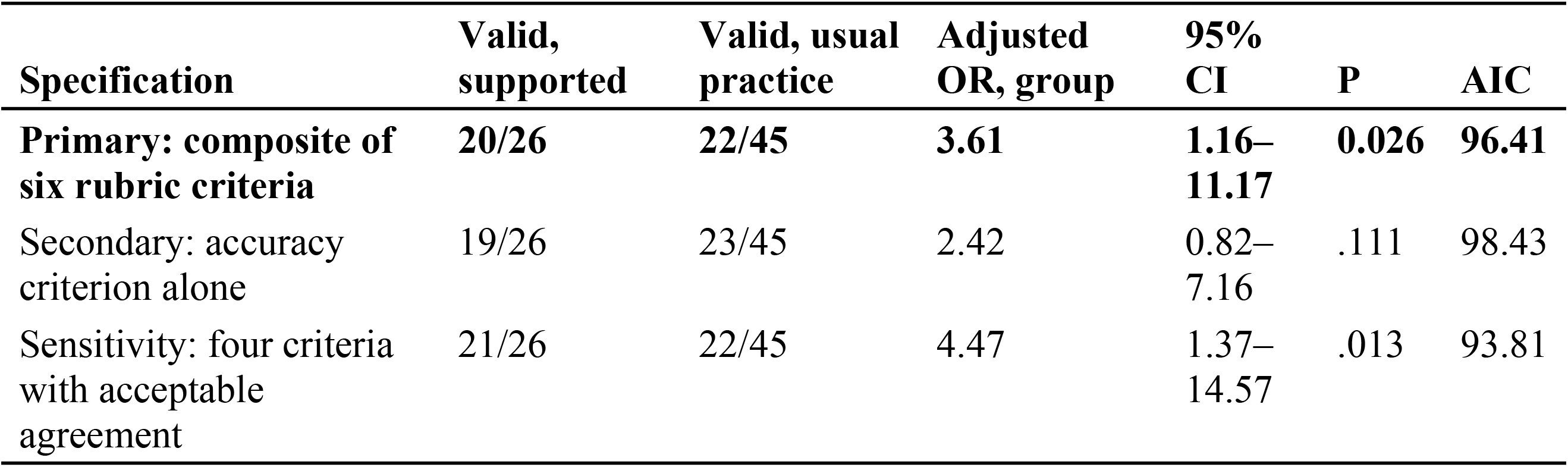
Multivariable logistic regression models for response validity.

All models adjust for academic degree, which was not independently associated with the outcome in any specification (primary model adjusted OR 1.12, 95% CI 0.39–3.19; P = .832). The sensitivity model excludes demographic bias and bias toward nonstandard treatment, the two criteria with the lowest interrater agreement (S2 Appendix, Table S10); agreement on the remaining four was itself no better than moderate. McFadden pseudo R² was 0.059, 0.038, and 0.078 respectively. For the primary composite model, BIC was 103.19, Cox-Snell R² 0.076 and Nagelkerke R² 0.103, and the likelihood ratio test against the intercept-only model gave χ²(2) = 5.63, P = .060, so the joint contribution of group and academic degree falls just short of conventional significance even though group alone is significant, which is consistent with the limited precision of this sample. The Hosmer–Lemeshow test is not informative for these models: with two binary predictors the fitted probabilities take only four distinct values and the data collapse into three risk groups (χ²(1) = 0.01, P = .930), so the test has almost no power to detect misfit.

### Efficiency

Response time was computed from the platform’s combined duration field, following the method described in Methods. Of 167 responses with a recorded duration of zero, 133 had a finished timestamp that preceded the started timestamp; these 167 responses, along with any others missing or exceeding one hour, were excluded from the efficiency analysis, leaving 969 responses from 70 participants, 25 in the supported arm and 45 under usual practice. The excluded responses were not evenly distributed across arms, but the loss was concentrated in a minority of physicians. Counted by response, 109 of 416 (26.2%) in the supported arm had no usable duration against 58 of 720 (8.1%) under usual practice; counted by physician, which respects the clustering of 16 responses within each participant, 9 of 26 supported physicians and 8 of 45 under usual practice had any missing timing record (Fisher exact P = .150), and the per-physician proportion of missing records did not differ significantly (Mann–Whitney P = .058). The response-level contrast therefore overstates the evidence for differential loss, because it treats clustered observations as independent; the imbalance is real in direction but not established at the level at which the efficiency outcome is analyzed.

Median time per question was 125.2 seconds (IQR 80.4–172.0) in the supported arm and 128.8 seconds (IQR 76.4–197.1) under usual practice, an absolute difference of −3.6 seconds (−2.8%) that did not reach statistical significance (P = .695). Time per case followed the same pattern (median 500.7 seconds, IQR 321.8–688.0, versus 515.0 seconds, IQR 305.5–788.3; P = .695).

Self-reported search counts were recorded for 704 of the 1,136 responses, corresponding to 44 participants, 15 in the supported arm and 29 under usual practice. This measure was missing to a similar extent in both arms (42.3% versus 35.6% of participants; Fisher exact P = .618). Among those with records, the median number of searches per question was 0.94 (IQR 0.53–1.03) in the supported arm and 0.56 (IQR 0.19–1.00) under usual practice (P = .184). Given the extent of missingness and the self-reported nature of the measure, this comparison is exploratory (Table 5).

**Table 5.**
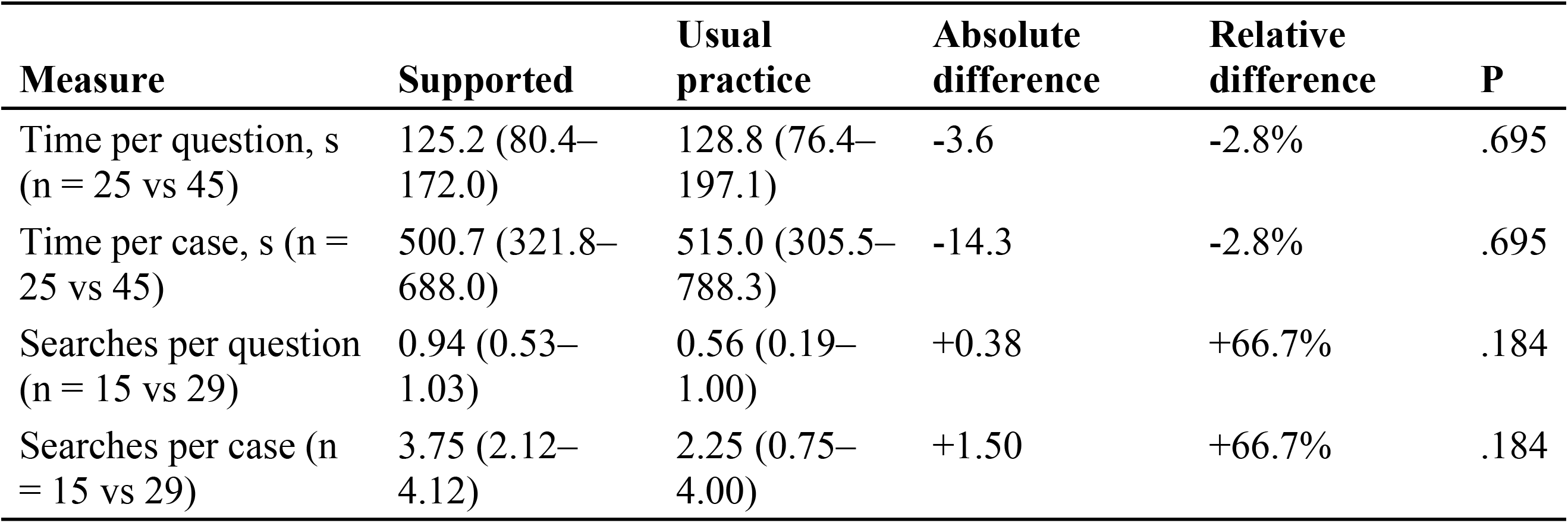
Response time and self-reported searches by group.

| Measure | Supported | Usual practice | Absolute difference | Relative difference | P |
| --- | --- | --- | --- | --- | --- |
| Time per question, s (n = 25 vs 45) | 125.2 (80.4–172.0) | 128.8 (76.4–197.1) | -3.6 | -2.8% | .695 |
| Time per case, s (n = 25 vs 45) | 500.7 (321.8–688.0) | 515.0 (305.5–788.3) | -14.3 | -2.8% | .695 |
| Searches per question (n = 15 vs 29) | 0.94 (0.53–1.03) | 0.56 (0.19–1.00) | +0.38 | +66.7% | .184 |
| Searches per case (n = 15 vs 29) | 3.75 (2.12–4.12) | 2.25 (0.75–4.00) | +1.50 | +66.7% | .184 |

Values are medians with interquartile ranges, compared with the Mann–Whitney U test. Absolute difference is supported minus usual practice. Values are computed from unrounded medians, so a per-case figure may differ in the last digit from the rounded per-question figure multiplied by four. Time is the platform’s combined per-response duration, retaining durations greater than zero and no longer than one hour; time per question is the mean of the valid durations and time per case is that value multiplied by four. Search counts were self-reported at the question level, computed the same way, and are missing for 432 of the 1,136 responses, so that analysis covers 44 participants and is exploratory. Both measures are reproducible from the deposited dataset with the accompanying script.

### Acceptability

Completing the acceptability questionnaire was a requirement to finish the exercise, yet a stored response exists for 25 of the 26 supported physicians. The single missing record is a data-capture failure, since the exercise could not be completed without submitting the questionnaire, and its cause could not be established retrospectively from the platform logs. Acceptability is therefore reported on 25 respondents. Overall acceptability was high, with a mean total score of 2.92 (95% CI 2.85–2.98). The system was rated useful for answering clinical questions (mean 2.88; 95% CI 2.76–3.00), and confidence in the truthfulness of its responses was the lowest-scoring item (mean 2.84; 95% CI 2.68–2.96). Likelihood of use in daily practice was high (mean 2.96; 95% CI 2.88– 3.00) and likelihood of recommending the system to colleagues reached the scale maximum (mean 3.00). Intervals are percentile bootstrap intervals over 20,000 resamples, which respect the bounds of the 1 to 3 scale. Item-level means are shown in Fig 2.

**Fig 2.**
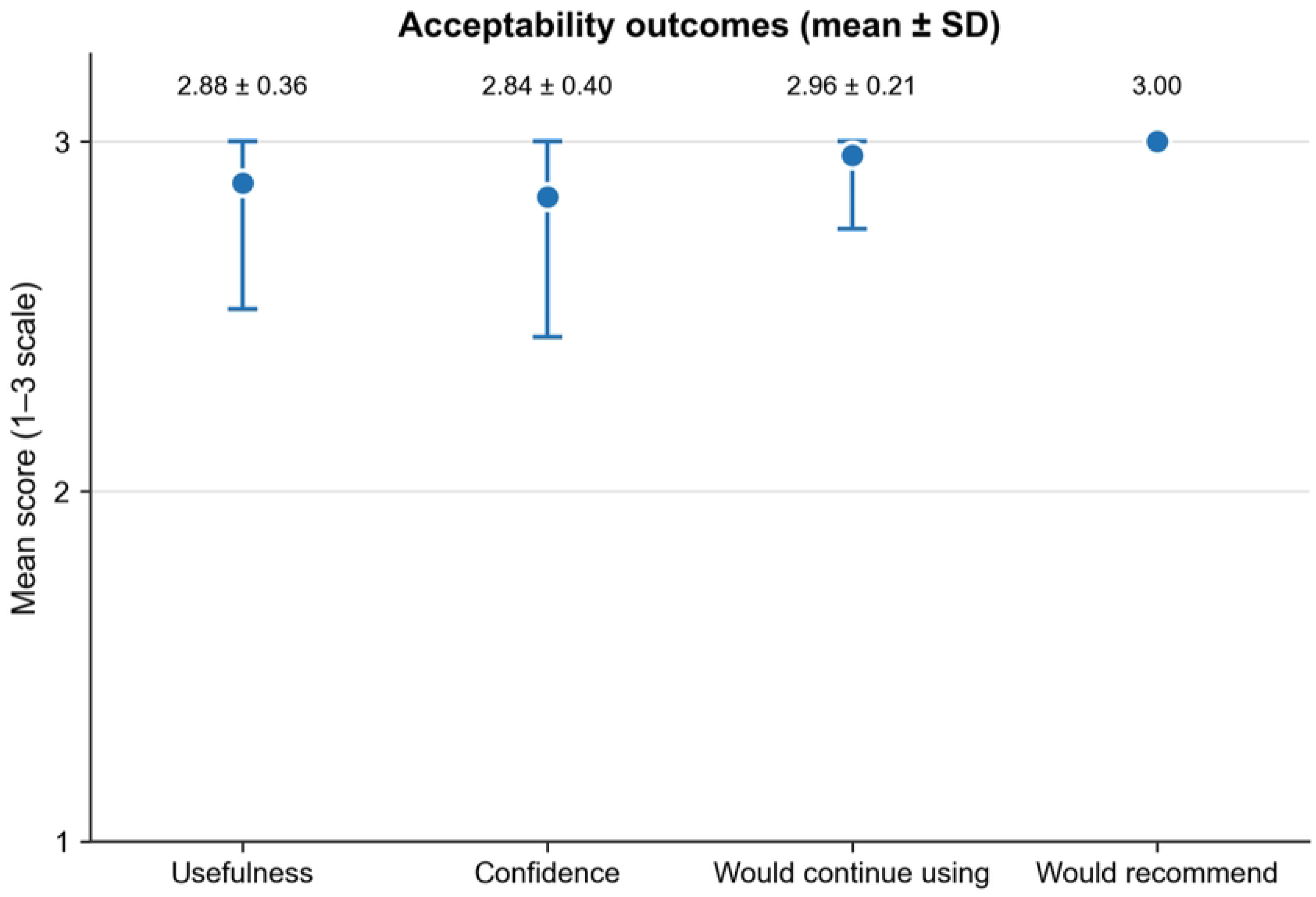
Acceptability of the conversational system among supported physicians. Mean score for each acceptability item on the 3-point scale, with standard deviations. Assessed only in the supported group (n = 25 with a stored response).

## Discussion

In this evaluator-blinded assessment, physicians supported by an evidence-traceable conversational system produced answers that independent specialists scored higher across all six rubric dimensions than answers produced under usual search practice, with a large effect size on total validity. The association persisted after adjustment for academic degree, with roughly threefold higher odds of meeting the validity threshold, at the physician level, in the supported group. Taken together, these results indicate a consistent association between system support and higher scored validity, estimated with limited precision.

How the outcome is operationalized changes the estimate, and the three specifications are reported in full for that reason. Modelling the composite of the six rubric criteria, which is the outcome the rubric was designed to capture, yields an adjusted odds ratio of 3.61. Restricting the outcome to a single criterion, accuracy, yields 2.42 with an interval crossing the null, and restricting it to the four criteria that achieved acceptable interrater agreement yields 4.47 with the best model fit. The direction is the same in all three, and the differences between them reflect how much information each specification retains: a single criterion discards five dimensions of clinical quality, while the composite preserves them. The three appear side by side because with 71 participants and wide confidence intervals the magnitude of the association is far less certain than its direction. This study was not registered prospectively, so these specifications cannot be checked against a prior analysis plan; they are reported in full so that readers can judge the stability of the finding for themselves.

The precision of these estimates is limited by design. With 71 participants and a low frequency of invalid responses in the supported group, confidence intervals are wide, and the lower bound of the primary estimate lies close to the null. The appropriate reading is that the direction of the association is consistent while its magnitude is not well determined, and that a confirmatory conclusion requires an adequately powered study rather than a reanalysis of these data.

The efficiency outcomes support no conclusion in either direction. Measured response time was nearly identical between arms (125.2 versus 128.8 seconds per question), but the records that produced those numbers were lost unevenly, in 26.2% of the supported arm’s responses against 8.1% under usual practice, so an equal measured time is as consistent with a real difference masked by selective loss as with no difference at all. An earlier analysis of these data found a larger apparent advantage for the supported arm; that estimate depended on treating a zero value in the platform’s combined timing field as a genuine instantaneous answer, and did not survive excluding those zeros once a substantial share of them proved to be inconsistent timestamp records, a distinction only detectable by cross-checking the raw timestamps. Search counts were missing to a similar extent in both arms, 42.3% against 35.6%, so that comparison is not distorted in the same way, but it is self-reported and available for fewer than two thirds of responses, which leaves it underpowered. What these data establish about efficiency is a measurement requirement rather than a result: response time needs a single, reliably captured, per-question field recorded server-side, which a study designed to estimate this outcome should adopt from the outset.

Acceptability was high across all items, with confidence in the truthfulness of responses scoring lowest among them. Physicians’ stated confidence in artificial-intelligence support does not necessarily track its measured reliability, which is the reason mechanisms that let clinicians calibrate that confidence have been proposed [32]. It reinforces the argument for provenance: systems that expose the literature behind each answer allow clinicians to calibrate trust by inspection rather than by assumption, which is precisely the property that most benchmark evaluations do not test.

### Comparison with existing evidence

These findings sit alongside a small body of work evaluating conversational systems with practicing clinicians [33,34]. Goh and colleagues found no significant difference in diagnostic reasoning between physicians with and without access to a large language model, attributing part of the null result to sample size and to a narrower outcome focused on diagnosis [14]. The present study differs in scope, assessing clinical validity across accuracy, guideline concordance, safety, timeliness, and bias rather than diagnostic accuracy alone, and on that composite it did detect a significant adjusted association. When the outcome here is narrowed to a single accuracy criterion, closer to what Goh and colleagues measured, the association also loses significance. That criterion still separates the arms when scores are compared directly (Table 3); it is the dichotomization that removes the contrast, because 24 of the 71 physicians sit exactly at the 2.5 threshold on that single criterion and are classified as valid under the inclusive rule. The contrast suggests that the breadth of the outcome may determine whether a benefit is detectable at these sample sizes, and that evaluations restricted to diagnostic accuracy may miss gains in other dimensions of clinical quality. Given the width of the confidence intervals in both studies, this remains a hypothesis for larger evaluations.

The evaluation instrument used here responds to a documented gap. The frameworks reviewed in the Introduction broadened the evaluative frame without producing a reference standard, and their linkage to clinical outcomes remains untested. The six-dimension rubric applied here is deliberately simple, was applied by domain specialists in their own areas, and is reported in full so that others can reuse or contest it. Its two weakest dimensions, demographic bias and bias toward nonstandard treatment, showed poor interrater agreement and are reported as exploratory, which is itself informative about which dimensions of clinical quality can be scored reliably by human experts with brief training.

### Relevance for Latin American health systems

External evaluations of clinical artificial intelligence conducted outside high-income settings remain uncommon, even though the constraints that make evidence retrieval difficult are more acute where specialist access is limited and clinical workload is high [10,25]. This study contributes evidence generated with practicing physicians in that context, using cases drawn from the four basic specialties available in most secondary-level referral hospitals in Colombia. That framing also bounds the claim: the results describe responses to four simulated clinical cases, including acute presentations, not performance during actual outpatient, emergency, or critical care.

### Strengths and limitations

The study’s principal strengths are its evaluator-blinded design with randomized invitation, applied with practicing physicians, and independent human scoring by domain specialists, which supports the clinical interpretability of the results. An external evaluator unaffiliated with the developer independently replicated the accuracy-based analysis, reported here as the secondary specification; the composite specification was designated primary after that audit and was not re-audited. Two specialists assessed each response in their own area, and the analysis used conventional statistical methods that clinical readers can scrutinize. The analytical matrices and reproduction scripts accompany this article.

Several limitations qualify these findings. The sample was a convenience sample without an a priori power calculation, and at 71 completing participants it yields wide confidence intervals and limited power to detect modest effects; the results therefore reflect performance under conditions of full completion. Although allocation was randomized at invitation, the analyzed sample is not a randomized sample. Only 202 physicians started, out of 8,891 contacted in the second wave and 221 registered in the first, and participation was itself unbalanced between arms, so selection operated before any outcome was measured. Among those who started, non-completion was substantial in both arms and higher under usual practice, where 99 of 144 (68.8%) did not complete, than in the supported arm, where 32 of 58 (55.2%) did not. The design is therefore better described as a randomized allocation followed by a complete-case analysis of volunteers than as a randomized comparison of allocated groups. Because the arm was disclosed in the invitation, assignment could influence both who started and who finished, so the selection operated after randomization and with knowledge of it. Non-completion in an asynchronous digital study is plausibly associated with lower technological affinity or lower motivation, so both analyzed groups are likely to over-represent physicians comfortable with digital tools, and neither can be treated as a random subset of those assigned.

The compensation compounds this in a way that is specific to this study: participants were offered premium access to the evaluated system itself, granted only on completing all sixteen responses. That reward is worth more to physicians who value such tools, and it was contingent on precisely the behavior that defines the analyzed sample. The reward plausibly pushes in the same direction as the observed difference in non-completion, which was lower in the supported arm, and cannot be separated from the effect of the assigned strategy with these data. A second asymmetry works the other way. Access in the supported arm expired automatically after two weeks without queries, so a physician in that arm who paused for longer lost the tool the exercise required, a barrier to finishing with no counterpart under usual practice, where the sources remained available throughout. The size of this effect is unknown, since the platform’s only record was of participation; what can be said is that the supported arm completed more often despite it.

The arms differed in academic degree from the moment physicians decided to take part, before any attrition: among all who started, 10 of 58 in the supported arm were specialists against 57 of 144 under usual practice (17.2% versus 39.6%, Fisher exact P = .003), and the difference persists among the analyzed completers (χ² = 4.96, P = .026); academic degree was retained in the model to control for residual confounding and was not independently associated with the outcome in any specification. The direction of the resulting bias is not determined by these data. The comparison group retained proportionally more specialists, but adjustment for academic degree cannot exclude selection on unmeasured characteristics or fully repair non-random loss. This remains the main threat to the internal validity of the comparison. Unequal group sizes alone do not invalidate the fitted models; here, however, the imbalance arose through voluntary participation and completion after allocation, so accommodating unequal sizes does not remove the associated selection bias.

Participant credentials were self-reported at registration and were not verified against a licensing registry, so the professional composition of the sample rests on what participants declared. The sampling frame compounds this. Neither recruitment wave drew on a population register: the first was an open call on social media and professional messaging groups, and the second was the developer’s own registry of physicians who had previously signed up for its educational activities or mailing list. Physicians on that registry had already shown an interest in clinical artificial intelligence before the study existed, which is the population most likely to engage productively with the tool under evaluation. The analyzed sample therefore skews young and digitally comfortable by construction, not only by attrition, and generalization to physicians with different levels of digital familiarity would require recruitment from a frame independent of the developer. That the second wave was undertaken because the first yielded too few completed responses also means the analyzed sample mixes two recruitment frames, and the study cannot say whether the two behave alike.

Clinical scenarios were restricted to four cases from core specialties, which bounds extrapolation to other cases and to clinical practice. The 3-point rubric, while standardized, offers limited granularity and produced scores clustered near the maximum. Ceiling effects and dichotomization discard information, but these data do not establish whether the resulting odds ratio is attenuated or exaggerated. Future evaluations should validate a more granular instrument and prespecify the outcome scale and analysis. One category ordering is open to question: for alignment with medical consensus, a response for which no consensus exists was scored 1 and a response opposed to consensus was scored 2, so a response contradicting established consensus ranks above one addressing a genuinely unsettled question. That ordering is difficult to defend clinically, and it was fixed before scoring began, so it cannot be corrected retrospectively. Re-scoring that criterion with the two levels exchanged leaves the finding intact, as reported in the Results, so the conclusion does not rest on that ordering, but a revised instrument should resolve it. The rubric was developed for this study by a small team that included employees of the developer and has not undergone formal psychometric validation, which is a potential source of systematic bias that the data alone cannot detect; its use is defensible given that no validated instrument for this purpose has been reported [18,19], and formal validation is a priority for subsequent work. The acceptability instrument was likewise developed for this study, has not been psychometrically validated, and is reported as descriptive user experience subject to the same ceiling effect, with three of its four items at or near the scale maximum.

The exact model configuration behind each answer cannot be fully reconstructed: the provider-side snapshot was not retained, and model metadata survives for only a minority of the study interactions. A reader cannot therefore reproduce the system exactly as participants encountered it, which limits the evaluation, though not its design, and one that a study of this kind should prevent by logging the model identifier with every response. Timing relied on client-recorded timestamps, with no server-side clock. A substantial share of records with a recorded time of exactly zero had a finished timestamp preceding the started timestamp, indicating that the client recorded the interval inconsistently; these records were excluded from the efficiency analysis. That exclusion was not balanced between arms: 109 of 416 responses (26.2%) in the supported arm lacked a usable duration, against 58 of 720 (8.1%) under usual practice, although the loss was concentrated in a minority of physicians and the per-physician difference does not reach significance (9 of 26 versus 8 of 45 with any missing record, P = .150). Because each physician’s time is summarized as the mean of the responses that do carry a duration, this affects how precisely each physician is characterized; the set of physicians entering the comparison is unchanged, and 70 of the 71 participants are represented, but supported physicians are summarized from fewer observations on average (a median of 16 responses in both arms, with a lower quartile of 7 in the supported arm against 16 under usual practice). If the responses that failed to record a duration differ systematically from those that did, it is not possible to say which way that would move the estimate. The efficiency comparison is therefore uninformative, and does not establish equivalence. Capturing timing server-side, per question, is a priority for a study designed to estimate this outcome precisely.

Search counts were self-reported and susceptible to recall and social-desirability bias, although missing to a similar extent in both arms, and future studies should capture search behavior automatically. Adherence to the assigned search strategy rested on participants’ compliance, and the source links recorded by the platform show departures in both arms. Under usual practice, where physicians were asked to rely on their customary sources, one physician linked to the evaluated system itself in 10 responses and another carried referral parameters from a general-purpose assistant in 3. In the supported arm, where the assigned system was to be the only source, three physicians linked to third-party assistants in 24 responses. Departures were therefore more frequent in the supported arm than in the comparison arm on this record, which removes the ground for treating the estimate as conservative: crossover in the comparison arm would narrow the observed difference, while third-party assistance in the supported arm could widen it, and the two cannot be netted against each other from these data. The record is also incomplete, since source links were captured only when participants supplied them and are present for a minority of responses, so these counts are lower bounds and their completeness may itself differ between arms. The contrast should be read as one between assigned strategies rather than between strictly separated exposures.

Evaluator blinding was supported by a platform that did not expose group assignment, and responses were presented in mixed order, but blinding was not formally verified. The study was not prospectively registered, for the reasons set out in the Methods. The consent document names the developer and the evaluated system, and both arms signed the same form, so physicians under usual practice knew they were serving as the comparator in an evaluation of that developer’s product. This is a plausible contributor to the higher non-completion under usual practice and to any difference in effort between arms, and its contribution cannot be disentangled from that of the assigned strategy here. Finally, most authors are employees of the organization that developed the evaluated system, a conflict mitigated but not eliminated by independent external replication of the accuracy-based secondary analysis and by public release of the analytical dataset.

These constraints define what the study can support: it establishes a reproducible evaluation approach and a directional signal in an underrepresented setting, and it identifies the specific design features, larger and more heterogeneous samples, stratification by training level, platform-captured efficiency measures, a wider scoring scale, and a psychometrically validated instrument, that a confirmatory study would need.

## Conclusions

Among practicing physicians in Latin America, answers produced with the support of an evidence-traceable conversational system were scored higher by blinded specialists than answers produced under usual search practice, and the association persisted after adjustment, with approximately threefold higher odds that a physician’s aggregate responses met the validity threshold. Response time and self-reported searches did not differ significantly, but timing records were unevenly missing between arms and that comparison should not be read as evidence of equivalence. These findings describe the validity of physicians’ answers under evaluation conditions, not clinical effectiveness or patient outcomes, and their precision is limited by sample size and differential attrition. The study contributes evaluation evidence from a setting underrepresented in the clinical artificial intelligence literature, together with a transparent six-dimension rubric and a reproducible analytical package that establish a basis for adequately powered confirmatory evaluation.

## Acknowledgments

We thank the specialists who contributed their expertise: Rafael Eduardo Arraut Gámez, Lina María Herrera Agudelo, Juan Nicolás Dallos Ferrerosa, Eder Donadoni Varela Macías, Germán Andrés Somoyar Duarte, Santiago José Morón Serrano, María Clara Mendoza Arango, and Juliana Muñoz Restrepo. We also thank Michael Andrés García Rivera for the independent external methodological review, including the reproducibility audit, and Dr Johan Morales Barrientos and Entrenarte S.A.S for their role in developing the clinical cases.

## Declaration of generative artificial intelligence use

During preparation of this manuscript the authors used DeepL for translation and language refinement, and an artificial intelligence coding assistant to support manuscript organization and formatting. All content was subsequently reviewed, edited, and approved by the authors, who take full responsibility for the integrity, accuracy, and originality of the published work.

## Data availability

The de-identified analytical matrices underlying the reported results, together with the scripts that regenerate them, are provided as Supporting Information. They cover the analyses of the 71 physicians included in the study: the validity comparisons of Table 3, the three logistic specifications of Table 4, the efficiency comparisons of Table 5 and the acceptability results. All 202 physicians who started the study provided informed consent at registration (S2 Appendix, Material S14a). The comparisons of completers and non-completers reported in S2 Appendix draw on the wider recruitment records of all 202; these records are not deposited in full because they include contact information collected for study administration, and are available from the corresponding author on reasonable request.

## Author contributions (CRediT)

**Natalia Castaño-Villegas:** Conceptualization, Formal analysis, Investigation, Data curation, Methodology, Validation, Writing – original draft. **Katherine Monsalve:** Methodology, Formal analysis, Writing – review and editing. **María Camila Villa:** Software, Writing – review and editing. **Oscar Iván Quirós Gómez:** Methodology, Writing – review and editing. **Laura Velásquez:** Resources, Supervision, Funding acquisition, Writing – review and editing. **José Zea:** Resources, Supervision, Writing – review and editing.

## Abbreviations

AIC: Akaike information criterion.
BIC: Bayesian information criterion.
CI: confidence interval.
CRediT: Contributor Roles Taxonomy.
IQR: interquartile range.
OR: odds ratio.

## Supporting information

**S1 Appendix. Data dictionary and reproduction guide.** Describes the deposited dataset, the construction of the outcome, and the scripts that regenerate Tables 3, 4 and 5.

**S2 Appendix. Supporting tables, clinical cases and consent documents.** Contains the supplementary tables referenced in the text, the four clinical cases with their questions, and the informed consent documents. Its analyses correspond to the secondary specification.

**S3 Data. De-identified analytical matrices and reproduction scripts (S3_Data.zip).** Contains the two de-identified matrices (one row per physician and one row per question-answer pair), the two scripts that regenerate the reported values from them, and a file describing how each variable was built.

## Notes

### Competing Interest Statement

Five of the six authors (N.C.V., K.M., M.C.V., L.V., J.Z.) are employees of Arkangel AI, the organization that developed the system evaluated in this study. O.I.Q.G. is affiliated with Universidad CES and declares no competing interests related to this work. To mitigate this conflict, an external evaluator unaffiliated with Arkangel AI independently replicated the accuracy-based analysis, reported in the manuscript as the secondary specification, and verified its reproducibility. The composite specification reported as primary was designated after that audit and has not been independently re-audited. The authors affiliated with Arkangel AI declare no financial competing interests beyond their employment. This does not alter the authors' adherence to PLOS ONE policies on sharing data and materials.

### Clinical Trial

N/A. This study randomly assigned practicing physicians to two information-search strategies and measured the quality of their answers to simulated clinical vignettes. All outcomes were measured at the level of the participating physicians, and no health outcomes of patients were assessed, so under the ICMJE definition the study does not meet the criteria of a clinical trial requiring registration. The reasoning is set out in the Methods section of the manuscript.

### Author Declarations

The study was approved by the Institutional Committee for Research Ethics in Humans (Comité Institucional de Ética en Investigación en Humanos) of Universidad CES, Medellín, Colombia, under project code Ae-1440, and was classified as risk-free research under Colombian Resolution 8430 of 1993. The protocol had been approved by the Research and Innovation Committee of the Faculty of Medicine of the same university on 17 September 2025, under code Acta321Proy012, before data collection began. All participating physicians and specialist evaluators gave electronic informed consent before taking part.

